# Multi-Stage Group Testing Improves Efficiency of Large-Scale COVID-19 Screening

**DOI:** 10.1101/2020.04.10.20061176

**Authors:** JN Eberhardt, NP Breuckmann, CS Eberhardt

## Abstract

**Background:** SARS-CoV-2 test kits are in critical shortage in many countries. This limits large-scale population testing and hinders the effort to identify and isolate infected individuals.

**Objectives:** Herein, we developed and evaluated multi-stage group testing schemes that test samples in groups of various pool sizes in multiple stages. Through this approach, groups of negative samples can be eliminated with a single test, avoiding the need for individual testing and achieving considerable savings of resources.

**Study design:** We designed and parameterized various multi-stage testing schemes and compared their efficiency at different prevalence rates using computer simulations.

**Results:** We found that three-stage testing schemes with pool sizes of maximum 16 samples can test up to three and seven times as many individuals with the same number of test kits for prevalence rates of around 5% and 1%, respectively. We propose an adaptive approach, where the optimal testing scheme is selected based on the expected prevalence rate.

**Conclusion:** These group testing schemes could lead to a major reduction in the number of testing kits required and help improve large-scale population testing in general and in the context of the current COVID-19 pandemic.

## Background

The COVID-19 pandemic is caused by the virus SARS-CoV-2 and despite drastic measures taken to limit disease dissemination, this pandemic will lead to a substantial death toll and an unforeseeable impact on health-care systems and the world economy. This shows the need for accurate data on prevalence to better inform political and public health decision making and to broadly identify infected individuals. So far, 1.6 million cases and over 100,000 deaths have been reported world-wide ([1], 10.4.2020). However, true prevalence rates are unknown as in most countries large-scale population testing has still not been introduced. Tests are often restricted to specific groups of populations, such as healthcare workers, individuals with known SARS-CoV-2 exposure, COVID-19 symptoms or with risk factors for severe diseases.

One reason for the limited testing is the shortage of PCR testing reagents. This could be overcome by group testing, a method first suggested by Dorfman for testing large populations of U.S. soldiers for syphilis [2]. The idea of group testing involves the division of the population into small groups. For each group, a combined sample (*‘pool’*) of its members is created and tested. If the pool tests negative, it can be concluded that all group members are negative and no individual tests will be required. If the pool tests positive, further tests will have to be performed to determine which group member(s) are positive. One ad-hoc model of group testing, using pools of up to 10 samples, has recently been applied for SARS-CoV-2 PCR [3]. More refined variants of group testing are for example used in HIV screening [3] [4]; but these pool sizes seem to exceed the sensitivity of current SARS-CoV-2 testing methods [5].

Herein, we develop various group testing schemes and evaluate their resource efficiency for different prevalence rates. We define models that are presumed practical for SARS-CoV-2 testing applications (small pool sizes, small number of steps) and best suited to minimize the number of tests necessary.

## Objectives

We developed and evaluated multi-stage group testing schemes that test samples in groups of various pool sizes in multiple stages. Through this approach, groups of negative samples can be eliminated with a single test, avoiding the need for individual testing and achieving considerable savings of resources.

## Study design

### Design of testing schemes

Multi-Stage group testing schemes *PNSk*(“pool size *N, k*stages”) were designed on the basis of two integers (divisor) and *k* (number of stages). The initial pool size is *N* = *x*^*k-*1^, which is divided by in each subsequent stage, resulting in pool sizes *x*^*k-*1^, *x*^*k-*2^,…, *x*^0^ = 1in stages1,2, …, *k*. Two-stage group testing applies this construction with setting *k* = 2.

### Evaluation of testing schemes

To compare the performance of group testing schemes, we defined a quantity called *improvement factor*. Mathematically, the improvement factor is the ratio of the population size and the expected value of the number of tests performed by the scheme. In other words, it is the average number of samples that can be tested with a single test, when the scheme is applied to a large population. Importantly, the improvement factor depends on the prevalence rate *p*.

Improvement factors of two-stage testing schemes *PNS2* were calculated using the formula *N*/(1 + *N* - *N*(1 - *p*)^*N*^))[4]. To handle multi-stage testing schemes, a PYTHON program was written by us. Additionally, PYTHON was used to implement a Monte-Carlo statistical method that performs multi-stage and matrix group testing schemes on 1M randomly generated groups of samples and averages the improvement factor over all groups. The two methods were compared and found to be in agreement with one another.

The improvement factors for all two-/multi-stage schemes with pool sizes up to 10,000 and for the (8×12) matrix scheme were calculated with the above methods for all prevalence rates *p* between 0% and 30% in steps of 0.05%. PYTHON was used to determine the optimal testing scheme amongst these examples. MATPLOTLIB was used to plot heatmaps visualizing the results.

We presume that schemes are clinically feasible if their pool size is less or equal than 16 and their number of stages is less or equal than 4.

A selection of presumed clinically feasible and optimal multi-stage schemes *P3S3, P9S3, P4S2* and *P16S3* was made. Also, the schemes *P32S2, P10S2* and the matrix scheme were considered as they appeared in earlier literature [6,7]. MATPLOTLIB was used to plot their improvement factors for prevalence rates between 0% and 30%. Data for prevalence rates over 30% was not plotted, since all testing schemes perform worse than individual testing in these cases.

## Results

### Design of group testing schemes

We designed group testing schemes with the goal of testing large numbers of samples more efficiently. Samples are not tested individually from the start but rather arranged into groups (‘pools’) and then tested together. All samples in pools that are tested negative must be negative and no individual testing is needed. All samples in pools that are tested positive are further processed according to the design of the testing scheme.

A commonly used approach is two-stage testing [2], where pools containing for example 3 individual samples (P3, “pool of 3”) are tested first, and in a second stage (S2, “2 stages”) samples in positive pools are tested individually (Fig 1A).

**Figure 1:**
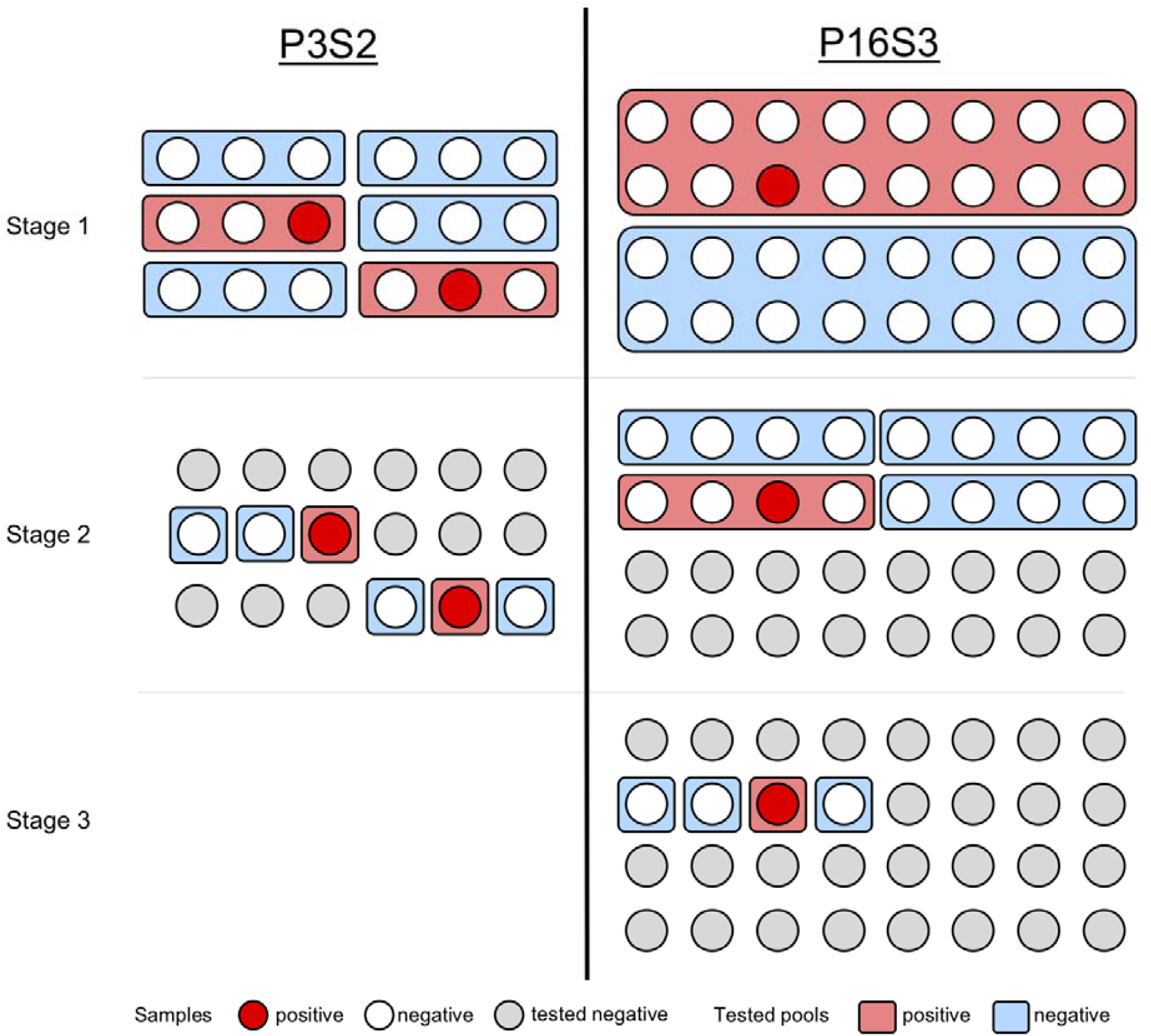
Schematic visualization of different group testing approaches. Scheme *P3S2* (left) is applied to 18 samples (circles) with 16 negatives (white) and 2 positives (red) samples. The spatial arrangement of the tests is irrelevant. Stage 1: 6 groups of 3 samples each are combined into pools (rectangles) and tested (blue for negative, red for positive). Stage 2: all samples belonging to a negative pool are considered negative and not further tested (grey). All samples from positive pools are tested individually. In total, 18 samples were tested with 12 tests (1.5 samples per test). With lower prevalence rates, *P3S2* can, on average, test up to 3 samples with 1 test. Scheme *P16S3* (right) is applied to 32 samples, one of which is positive. Stage 1: 2 groups of 16 samples are pooled and tested. Stage 2: All samples in the negative group must be negative and are hence not tested further. Samples in the positive group are pooled into 4 subgroups of 4 samples and each pool is tested. Stage 3: The remaining 4 samples in the one positive pool are tested individually. In total, 32 samples were tested with 10 tests (3.2 samples per test). With lower prevalence rates, *P16S3* can, on average, test up to 16 samples with 1 test.

The resource efficiency of group testing stems from the fact that for low prevalence rates it is likely that a group of samples will not contain a positive sample and thus negative samples are eliminated in groups.

Group testing schemes can be refined in various ways. We expanded the design to *multi-stage testing schemes*. Here, pools that are tested positive are split in multiple stages into smaller pools before eventually performing individual tests. We used integer powers *x*^*k*^(e.g. 4^*k*^: 1, 4, 16, …) of pool sizes and divided them by the fixed number *x* (divisor) after each stage. For example, a testing scheme that we call *P16S3* (“pool of 16, 3 stages”) has the divisor *x*= 4, meaning that pool sizes are divided by 4 at each stage: It uses pools of 16 samples in the first stage, pools of 4 samples in the second stage and individual testing in the third stage (Fig 1B).

There are obvious variants, which are more complicated, such as using different divisors at each stage, but we did not consider these here.

### Evaluation of different two- and multi-stage group testing schemes

To evaluate and compare the performance of different group testing schemes, we defined the *improvement factor* compared to individual testing. The improvement factor describes how many more samples can be tested using the same (limited) number of ressources. Simplistically speaking, an improvement factor of 10 indicates that 20,000 tests are sufficient to test an average population of 200,000 individuals.

The improvement factor of each scheme depends on the prevalence rate. A calculation shows that for a prevalence rate *p* and a pool size *N* the improvement factor for two-stage testing is *N*/(1 + *N* − *N*(1 − *p*)^*N*^). Hence, if the prevalence rate approaches 0, the improvement factor approaches the pool size N. Multi-stage testing schemes have the same asymptotic behaviour although explicit formulas for their improvement factor become more complicated.

We next compared the improvement factors of each testing scheme assuming different prevalence rates up to 30%. Results for selected prevalence rates (1%, 7.5% and 20%) are visualised in Figure 2.

**Figure 2:**
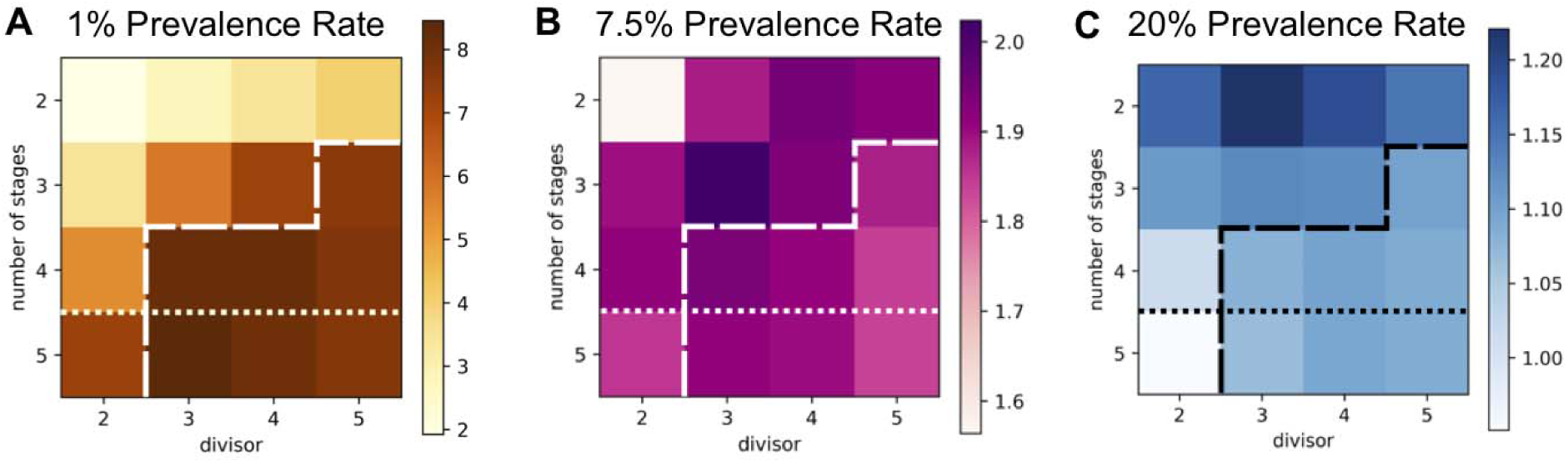
Performance of various multi-stage schemes under prevalence rates of 1%, 7.5% and 20%. Each square represents a multi-stage scheme *PNSk* with pool size *N* = *x*^*k-*1^, divisor (x-axis) and *k* stages (y-axis). Color intensity represents the improvement factor, i.e. the average number of subjects tested with a single test (darker is higher). Dashed lines indicate the cut-off with respect to the maximal initial pool size of 16. Dotted lines indicate the cut-off with respect to the maximal number of stages of 4. **A:** For 1% prevalence rate, among all schemes, *P81S5* performs best (improvement factor 8.4). Among schemes with a pool size limited by 16, *P16S5* performs best (improvement factor 7.3). Finally, among schemes which are additionally limited to less than 5 stages, *P16S3* performs best (improvement factor 7.1) testing 7.1 subjects with just one test, while still being practical. **B:** For 7.5% prevalence rate, *P9S3* performs best (improvement factor 2). **C:** For a 20% prevalence rate, it is preferable to use a low number of stages. *P3S3* performs best (improvement factor 1.22).

The data showed that group testing is more efficient than individual testing for prevalence rates under 30%. As expected, large pool sizes and more stages are preferable for lower prevalence rates, small pool sizes and fewer stages are preferable for higher prevalence rates, indicating that there is no group testing scheme that is optimal for all prevalence rates.

For prevalence rates under 12%, multi-stage schemes had higher improvement factors than two-stage schemes. For prevalence rates around 1%, simple three-stage schemes such as *P9S3* or *P16S3* (Fig 1B) yielded improvement factors of around 7.

At prevalence rates of around 1%, group testing schemes with very large pool sizes and many stages, for example *P81S5*, are optimal in the mathematical sense but clinically impractical for several reasons: First, PCR testing is not arbitrarily sensitive. Pooling positive with negative samples dilutes the positive samples, which increases the risk of false negative results. However, recent research indicates that pooling of 16 samples does not seem to substantially impact test sensitivity of SARS-Cov-2 PCR tests [5]. Second, the number of stages should be reasonable. Each additional stage increases the workload and the risk of human error. Furthermore, stages have to be processed sequentially, increasing the time to get the diagnosis. Therefore, we decreased complexity and restricted our further analysis to schemes with pool sizes up to 16 samples and a maximum of 4 stages.

### Performance of presumed clinically feasible group testing schemes

We determined the improvement factor of *P3S2, P9S3, P4S2* and *P16S4* for prevalence rates up to 30% (Figure 3 A+B) and compared results with previously described schemes, namely the two-stage testing schemes with pool sizes of 10 (*P10S2*, [6]), or 32 (*P32S2*, [5]). We also assessed the matrix testing scheme [7] using a matrix of 96 samples (12×8) that groups samples in rows (8 pools comprising 12 samples) and columns (12 pools comprising 8 samples) a method that has also been used for epitope mapping in immunology research application [8].

**Figure 3:**
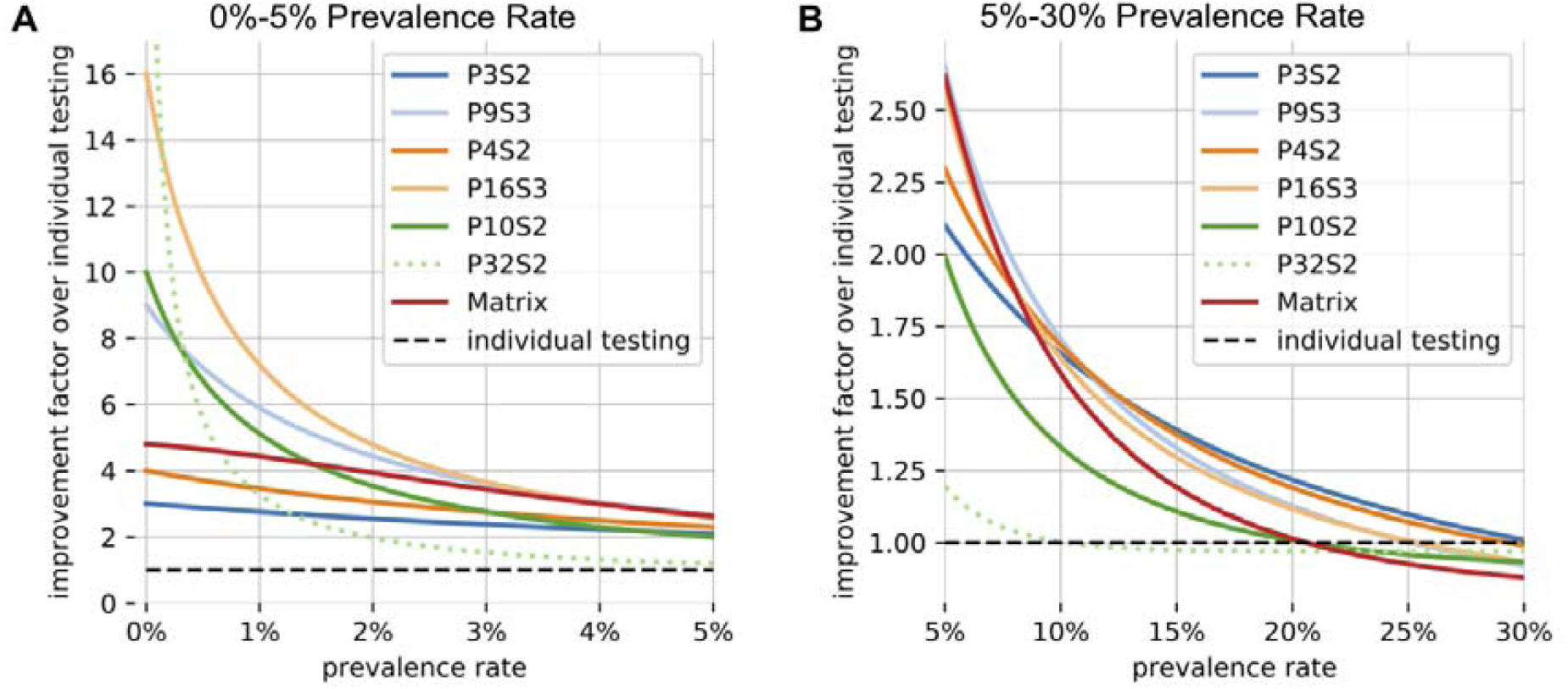
Improvement factors of different schemes for prevalence rates below 30%. **A:** Improvement factors of the different schemes for prevalence rates below 5%. For prevalence rates below 3.5% scheme *P16S3* is favorable, leading to an improvement factor of between 3-16-fold. Above 3.5% scheme *P9S3* becomes favorable, giving improvement factors of around 3-fold. Note that for very low prevalence rates the improvement factors of multi-level schemes converge towards the maximum pool size, making schemes such as *P10S1, P16S3* and *P32S2* highly efficient. *P32S2* is shown with a dashed line since its large maximum pool size may affect the reliability of the tests. For prevalence rates < 0.5% it rapidly converges towards an improvement factor of 32-fold. **B:** Improvement factors of the different schemes for prevalence rates between 5-30%. The data shows that for prevalence rates below 12% scheme *P9S3* gives the largest improvement rate, whereas above 12% scheme *P3S2* becomes favorable. For prevalence rates of 30% and above all schemes considered here do not offer an advantage over individual testing. Schemes with a large maximum pool size (*P10S2, P32S2, Matrix*) offer lower improvement rates and are hence unfavorable in this regime.

We summarized the optimal testing schemes for different prevalence rates (Table 1):

**Table 1:** Detailed summary of testing schemes and improvement factors for various prevalence rates. Schemes which are compatible with our desiderata for clinical practicability (pool size ≦ 16 and low number of stages ≦ 4) and their properties. We show which scheme has the best performance for any underlying prevalence rate.

| Scheme | Maximum pool size | Minimum number of samples | Number of stages | Best for prevalence rates of (Improvement factor) |
| --- | --- | --- | --- | --- |
| *Individual Testing* | 1 | 1 | 1 | 30% and above |
| *P3S2* | 3 | 3 | 2 | 12-30% (1.5-1) |
| *P9S3* | 9 | 9 | 3 | 3.5-12% (1.5-3.8) |
| *P4S2* | 4 | 4 | 2 | - |
| *P16S3* | 16 | 16 | 3 | 0.-3.5% (16-3.8) |
| *P10S2* | 10 | 10 | 2 | - |
| *Matrix* | 12 | 96 | 2 | - |

For low prevalence rates (0-3.5%), *P16S3* is optimal among all the schemes we consider feasible, with improvement factors between 16 down to 3.8. For medium prevalence rates (3.5-12%), it becomes advantageous to reduce the pool size to 9 and perform 3 stages (*P9S3*), giving an improvement factor from 3.8 down to 1.5. For high prevalence rates between 12% and 30%, the pool size should be further reduced to three samples with 2 stages (*P3S2*), yielding an improvement factor from 1.5 down to 1. Once the prevalence rate is 30% and above, individual testing should be performed.

In summary, we showed that among all presumed clinically feasible testing schemes multi-stage schemes are preferable to two-stage schemes for prevalence rates of under 12%. For prevalence rates up to 3.5% *P16S3* is preferable. From 3.5% up to 12% *P9S3* performs best. However, for prevalence rates above 12% the two-staged testing scheme *P3S2* is preferable.

## Discussion

We introduced *multi-stage group testing schemes* which are highly efficient methods to test large numbers of samples and assessed their *improvement factor* depending on different prevalence rates. We found that three-stage schemes performed optimally for prevalence rates up to 12% and that initial pool sizes of 16 were best for prevalence rates up to 3.5% (*P16S3*, improvement factor 16 to 3.8) and pools of 9 samples for rates between 3.5% and 12% (*P9S3*, improvement factor 3.8 to 1.5). For prevalence rates between 12 and 30%, two-stage testing with pools of 3 samples performed best (*P3S2*, improvement factor 1.5 to 1). This suggests that an adaptive approach is necessary where the scheme is chosen depending on the estimated prevalence rate. The multi-stage group testing schemes defined in this paper outperform other approaches to group testing in the literature [7].

The high efficiency of multi-stage group testing allows for large-scale testing of populations. To estimate the potential savings of tests we can use real-world data [1] (dated 10.04.2020). In order not to overestimate the prevalence rate, we considered data from countries which have performed large-scale population testing, namely South Korea and Germany. In South Korea a total of 503,051 people have been tested, of which 10,450 were positive. This gives an estimate for the underlying prevalence rate of 2%. At this prevalence rate the multi-stage testing scheme *P16S3* is optimal and would allow for testing *about five times* as many individuals with the same number of tests. In Germany, a total of 1,317,887 people were tested, of which 118,235 were positive. This gives a higher prevalence rate of around 9%. In this case the multi-stage scheme *P9S3* is optimal; it would enable the testing of an *additional 80%* of individuals (compared to only 30% improvement based on the *P10S2* scheme suggested in [6]). Note that these numbers have a selection bias, as individuals with COVID-19 symptoms were more likely to be tested. By introducing large-scale testing, established true prevalence rates will probably be lower, which would automatically yield even better improvement factors of the testing schemes.

Our analysis and predictions are *in silico* observations and have to be confirmed in real life. In particular, these testing schemes will have to be established in a similar fashion as other novel individual laboratory testings with the aim of assessing and limiting potential false positive or negative results. The practical feasibility of our methods is supported by observations of Yelin *et al*. which indicate that pooling up to 16 samples for SARS-Cov-2 PCR testing could potentially not decrease test sensitivity [5]. If further laboratory analyses show that the scheme *P16S3* is not implementable and that pool sizes of 16 samples increase the false negative rate significantly, the scheme *P9S3* presents a viable alternative. Here, improvement factors are comparable (7.2 vs. 6 for 1% prevalence rate) while almost halving the pool size.

However, it has been described that the viral load depends on the disease stage and is high during the early phase [9]. Thus, larger pool sizes could be used for this subpopulation. Additionally, individuals with higher and lower likelihood of infection could be combined into different pools to improve testing efficiency by choosing the appropriate testing scheme.

As these group-testing schemes are agnostic towards practical application, they can be used in different settings. In the course of the current COVID-19 pandemic and in view of the shortage of PCR testing kits, these multiple-stage schemes would allow for large-scale population testing and prevalence estimations. Also, efforts are made to determine sero-prevalence of anti-SARS-CoV-2 antibodies as potential markers of previous infection and immunity and broad population testing will soon become necessary. Here, multiple-stage testing could be employed and it has been shown for anti-HIV antibody testing that ELISA tests of pooled serum samples can be performed without a significant decrease in sensitivity or specificity [10].

In summary, we identified group testing schemes that are more efficient than individual testing methods most laboratories currently employ under different prevalence rates. These findings can have the potential to significantly increase the mass testing efficiency in the context of the current COVID-19 pandemic.

## Data Availability

The authors confirm that the data supporting the findings of this study are available within the article and its supplementary materials.

## Acknowledgements

We thank John Kwan for the careful reading of our manuscript and CoCalc for providing a remote collaborative programming platform.

## Notes

### Competing Interest Statement

The authors have declared no competing interest.

### Funding Statement

No external funding was received.

